# Polysubstance Injection and Smoking Trajectories of Unregulated Drug Use in the San Diego-Tijuana Border Region: A Latent Transition Analysis

**DOI:** 10.64898/2026.05.27.26354253

**Authors:** William H. Eger, Angela R. Bazzi, Erika L. Crable, Daniela Abramovitz, Alicia Harvey-Vera, Carlos F. Vera, M. Gudelia Rangel, Joseph R. Friedman, Eileen V. Pitpitan, Thomas L. Patterson, Steffanie A. Strathdee, Heather A. Pines

**Affiliations:** School of Social Work, San Diego State University, San Diego, California, USA; School of Medicine, University of California San Diego, La Jolla, California, USA; Herbert Wertheim School of Public Health, University of California San Diego, La Jolla, California, USA; Boston University School of Public Health, Boston, MA, USA; Fielding School of Public Health, University of California Los Angeles, Los Angeles, California, USA; Center for Health Policy Research, University of California Los Angeles, Los Angeles, California, USA; Department of Population Studies, Colegio de la Frontera Norte, Tijuana, Mexico; School of Public Health, San Diego State University, San Diego, California, USA

**Author notes:** Corresponding author: William H. Eger, MPH, PhD, Candidate, School of Social Work, San Diego State University, Address: 5500 Campanile Dr, San Diego, CA 92182. **Email addresses of authors:** ARB, ELC, DA, AHV, CFV, MGR, JRF, EVP, TLP, SAS, HAP.

**Keywords:** Fentanyl, Substance Abuse, Intravenous, Smoking, Methamphetamine, Drug Overdose, Harm Reduction

## Abstract

**Background and Aims:** The North American overdose crisis is increasingly characterized by complex polysubstance use alongside a transition from injecting to smoking unregulated opioids. However, transitions involving multiple substances remain understudied. We characterized longitudinal transitions in the route of administration and frequency of heroin, fentanyl, and methamphetamine use and examined whether these transitions differed by multilevel factors hypothesized to influence patterns of polysubstance use and routes of administration over time.

**Design:** People who inject drugs (PWID) enrolled in a cohort study completed baseline surveys (October 2020–2021) and three biannual follow-up visits (through April 2023).

**Setting:** San Diego, California, and Tijuana, Baja California.

**Participants:** Among 612 PWID, median age was 43 years; most were male (74%), Hispanic, Latino, or Mexican (72%), and San Diego residents (67%).

**Measurements:** Based on past six-month substance use behaviors reported at each visit, we categorized participants according to six indicators over time: low- (< weekly) and high-frequency (≥ weekly) smoking and injecting of heroin, fentanyl, and methamphetamine. We then used latent transition analysis (LTA) to identify distinct subgroups of participants with respect to these indicators at baseline and examine transitions between them over 18 months. We fit models with 2–5 subgroups, selecting the final model based on fit and interpretability and used multiple-groups LTA to examine differences in subgroup transitions by multilevel factors.

**Findings:** We identified four subgroups: Subgroup 1 (“*Heroin-Methamphetamine Polyroute*”), characterized by high-frequency heroin and methamphetamine smoking and injection, included 22% of participants at baseline but 0% at 18 months. Subgroup 2 (“*Methamphetamine-dominant Smoking*”), characterized by high-frequency methamphetamine smoking, accounted for 14% of participants at baseline and 18 months. Subgroup 3 (“*Fentanyl-Methamphetamine Smoking*”), characterized by high-frequency fentanyl and methamphetamine smoking, included 4% of participants at baseline and 21% at 18 months. Subgroup 4 (“*Heroin-dominant Injecting*”), characterized by high-frequency heroin injection, included 61% of participants at baseline and 65% at 18 months. Participants in Subgroup 1 primarily transitioned to Subgroups 3 and 4 over time. Larger increases in Subgroup 3 prevalence occurred for participants who, at baseline, experienced homelessness, resided in San Diego (vs. Tijuana), received syringes from a syringe services program, and overdosed in the past six months.

**Conclusions:** PWID in this region increasingly transitioned from high-frequency heroin and methamphetamine injection toward fentanyl and methamphetamine smoking, likely reflecting shifts in drug availability. Results highlight the need for multilevel interventions that address health harms resulting from polysubstance smoking alongside continued injection.

## INTRODUCTION

Coinciding with the Fourth Wave of the North American overdose crisis, defined by polysubstance use involving unregulated fentanyl and methamphetamine (1, 2), there have been substantial reductions in the prevalence of opioid injection and corresponding increases in opioid smoking (3–6). While the prevalence of methamphetamine injection also appears to have declined (4), this trend has received less attention. Transitions from injecting to smoking unregulated drugs are frequently framed as risk-reducing (7, 8); compared to injecting, smoking is associated with a lower risk of blood-borne infections, reduced incidence of skin and soft tissue infections, and avoidance of venous damage (9–11). However, uncertainties regarding health harms remain: smoking fentanyl and methamphetamine is increasingly implicated in overdose mortality (12, 13), and the shorter duration of effect associated with smoking may lead to higher-frequency use, potentially offsetting some perceived benefits of non-injection drug use.

Explanations for the observed shifts from injecting to smoking unregulated drugs have mostly focused on opioids and individual-level factors, including overdose risk perceptions, experiences of injection-related injury or infection, and difficulties with venous access (3, 7, 8). While important, it is unlikely that these factors fully explain the magnitude and geographic variability of observed shifts in routes of drug administration. For example, declines in injecting appear to be greater on the West Coast of North America than on the East Coast (5), even as recent reductions in overdose mortality have been more broadly distributed (14). This pattern suggests that additional multilevel factors may influence transitions from injecting to smoking.

Drug market transformations, long considered a structural factor (15), provide one critical lens through which recent shifts in routes of unregulated drug use may be understood. The introduction of unregulated fentanyl into the North American heroin supply occurred earlier and more abruptly in eastern regions, where fentanyl initially appeared as an unrecognized adulterant (16). In contrast, western drug markets experienced later but more rapid fentanyl penetration, often accompanied by overt marketing of fentanyl as a distinct product (17, 18). These regional differences in fentanyl perception and marketing may further explain the higher prevalence of fentanyl smoking on the West Coast, underscoring the role of place in shaping substance use. In addition, prior research on transitions in routes of unregulated drug administration has focused almost exclusively on opioids, despite methamphetamine being a commonly used unregulated substance on the West Coast of North America and a key driver of polysubstance use and overdose risk (19–21). Methamphetamine’s pharmacologic properties, patterns of co-use with fentanyl, and evolving forms in the unregulated drug market may produce distinct trajectories in routes of drug administration. Understanding whether and how people transition from injecting to smoking methamphetamine—and how these transitions intersect with opioid use—is a critical and understudied component of the current polysubstance overdose crisis (2).

The San Diego–Tijuana border region (SDTBR) represents an informative setting for examining transitions from injecting to smoking in the context of complex polysubstance use because of its drug market dynamics. As a binational drug market and major transit corridor for heroin, fentanyl, and methamphetamine entering North America (22), the region has experienced rapid shifts in drug availability, composition, and price (23). San Diego encountered widespread fentanyl penetration earlier than many other western cities in the United States (U.S.), followed by sharp increases in overdose mortality (24, 25). However, fentanyl appears to have penetrated the local drug supply in Tijuana more slowly, where it may not have been as overtly marketed (26). Despite growing recognition of population-level shifts away from injecting, little is known about how trajectories of polysubstance injecting and smoking evolve over time within individuals, or how these trajectories may vary by factors beyond the individual level. Studying transitions from injecting to smoking in the SDTBR offers an opportunity to better understand how these dynamics are shaped by time, geography, and drug markets conditions.

Identifying multilevel determinants of transitions from injecting to smoking can help inform the development and delivery of interventions to reduce infectious disease and overdose morbidity and mortality among people using opioids and stimulants in the fentanyl era (27). Latent transition analysis (LTA) offers an opportunity to fill current gaps as it can identify unobserved population subgroups and transitions between those subgroups based on observed patterns of behavior (28, 29). We conducted a LTA of heroin, fentanyl, and methamphetamine injecting and smoking among people who inject drugs (PWID) in the SDTBR to identify distinct subgroups of PWID with respect to their substance use patterns and transitions between those subgroups over time. Guided by the social ecological model (SEM), we further examined how transitions between subgroups differed by overdose history, access to syringe services programs (SSPs), housing status, and location of residence to understand how these multilevel factors may have influenced shifts from injecting to smoking unregulated drugs across North America (30).

## METHODS

### Data source

Data are from 612 PWID who participated in a longitudinal cohort study (*“La Frontera”*) conducted between October 2020 and April 2023 to examine the concurrent HIV, HCV, and drug overdose syndemic in the SDTBR. As previously described (31), *La Frontera* staff employed several evidence-based strategies to recruit PWID (32, 33) through a) research team-leased drop-in centers strategically located in areas frequented by PWID; b) street-based outreach conducted via mobile vans; and c) informal peer referrals through participants’ social networks (“snowball sampling”). Individuals ≥18 years of age, fluent in English and/or Spanish, living in San Diego or Tijuana, and who injected drugs in the past month were eligible. After providing written informed consent, participants completed one-hour, interviewer-administered surveys at baseline and follow-up visits every six months for 18 months in English or Spanish depending on their language preference. All research activities were approved by the Institutional Review Boards at the University of California, San Diego, and Universidad Xochicalco.

### Study measures

*Substance use indicators* capture participants’ self-reported substance use in the past six months. At each study visit, participants reported on their frequency of heroin, fentanyl, and methamphetamine use by route of administration via the following response options: *never, once per month or less, 2-3 days per month, once per week, 2-3 days per week, 4-6 days per week, once per day, 2-3 times per day,* and *4 times or more per day*. Based on this study’s objectives, we then created six binary substance use indicators to classify participants as engaging in *no- to low-frequency* (less than once per week) or *high-frequency* (at least once per week) i) heroin injection, ii) heroin smoking, iii) fentanyl injection, iv) fentanyl smoking, v) methamphetamine injection, and vi) methamphetamine smoking at each study visit. Substance use frequency cut-offs (*no- to low* vs. *high*) were informed by the distribution of baseline substance use behaviors and insights from concurrent in-depth qualitative interviews conducted with the cohort (Supplemental Table 1).

*Multilevel exposures of interest* were measured at baseline and included multilevel factors hypothesized to influence transitions from injecting to smoking heroin, fentanyl, and methamphetamine based on the SEM (30), and be most informative for the development and delivery of interventions focused on reducing infectious disease and overdose morbidity and mortality based on the literature (3, 34), qualitative evidence from this cohort, and study team expertise. Exposures included location of residence (San Diego or Tijuana; *structural*), and past six-month housing status (unsheltered homelessness, sheltered homelessness, or not experiencing homelessness; *structural*), receipt of syringes from an SSP (*community*), and overdose history (*individual*). For analysis, we collapsed housing status into a binary variable representing whether participants had or had not experienced (unsheltered or sheltered) homelessness.

*Sociodemographic characteristics* were measured at baseline and included age in years, sex assigned at birth (female or male), ethnicity (non-Hispanic/Latino/Mexican or Hispanic/Latino/Mexican), average monthly income (<500 or ≥500 United States dollars [USD]), highest level of education completed (at least secondary school or more than secondary school), and incarceration history (never or ever in jail/prison).

### Statistical analysis

To characterize subgroups of participants that exhibited distinct patterns in their frequency of injecting and smoking of heroin, fentanyl, and methamphetamine, and transitions between those subgroups over time, we leveraged substance use behaviors reported by participants over 18 months (i.e., at enrollment [first enrollment date: October 28, 2020] and at their follow-up visits every six months thereafter [through April 27, 2023]) in a LTA conducted using PROC LTA in SAS v9.4 (28, 35). PROC LTA uses an expectation-maximization algorithm, which assumes data are missing at random and that differences between observed and missing indicators can be explained by available relationships within the observed data (35). Therefore, participants who did not return for follow-up were not excluded from analyses; rather, their observed information was used to probabilistically estimate their parameters (36). Characteristics of participants who missed at least one follow-up visit versus those who did not are included in Supplemental Table 2. In addition, to understand the extent to which our findings reflect subgroup transitions over time among those with complete follow-up data, we conducted a sensitivity analysis excluding participants who missed at least one follow-up visit (n=166).

We fit a series of LTA models with two to five subgroups, constraining item-response probabilities to be equal across time, to identify the number of subgroups that provided the optimal balance of fit and parsimony (28, 29). Consistent with recommended approaches for LTA (37, 38), model selection was guided by multiple complementary fit indices, including the likelihood-ratio G^2^ statistic, Akaike Information Criterion (AIC) and Bayesian Information Criterion (BIC) (39, 40), as well as substantive interpretability of the identified subgroups. Using our final LTA model, we estimated i) item-response probabilities for each subgroup at baseline (i.e., the probability of endorsing each substance use indicator conditional on subgroup membership), ii) subgroup membership prevalences at each time point, and iii) transition probabilities between subgroups across adjacent six-month time points.

Next, to understand the extent to which subgroup transitions may differ across groups defined by strata of our multilevel exposures of interest, we fit separate multiple-groups LTA models for each exposure, fixing the number of subgroups to that identified in our final model. We first assessed measurement invariance by fitting two models for each exposure: one with item-response probabilities estimated freely, and one with these probabilities constrained to be equal across groups defined by strata of the exposure. We compared the fit of these models using a likelihood ratio test (G^2^ difference test) (35). Based on these results, we specified a common measurement model for each multiple-groups LTA model and constrained item-response probabilities to be equal, while allowing transition probabilities to vary across groups (29). As the multiple-groups LTA model for ‘location of residence’ did not converge, we used the posterior probabilities from our final LTA model to assign participants to one subgroup at each time point for which they had the highest posterior probability of membership (38, 41) and then examined subgroup prevalences by ‘location of residence’ over time.

## RESULTS

### Sample description

Among 612 participants at baseline (Table 1), 77% reported high frequency heroin injecting. Approximately one-third reported high frequency methamphetamine injecting (36%) and 52% reported high frequency methamphetamine smoking. Less than one-quarter reported high frequency heroin smoking (18%) and high frequency fentanyl injecting (16%) and smoking (11%). Two-thirds were San Diego residents (67%), and, in the past six months, 43% experienced unsheltered homelessness, 37% received syringes from an SSP, and 16% experienced an overdose. Participants had a median age of 43.0 years (IQR: 35.0, 52.0) and most were male (74%), Hispanic, Latino, or Mexican (72%), had a monthly income less than $500 (55%), completed more than secondary education (57%), and were ever incarcerated (65%).

**Table 1.**
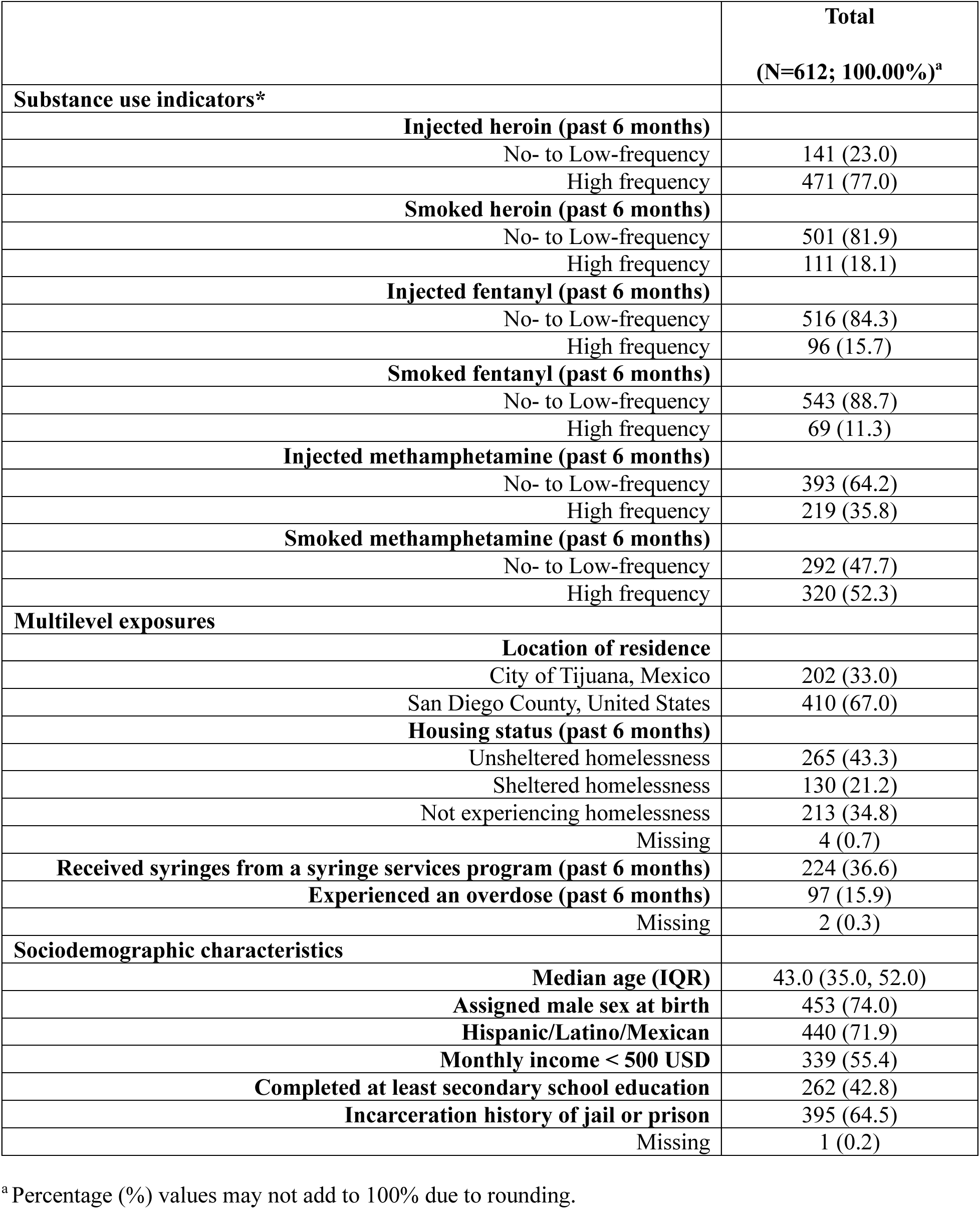
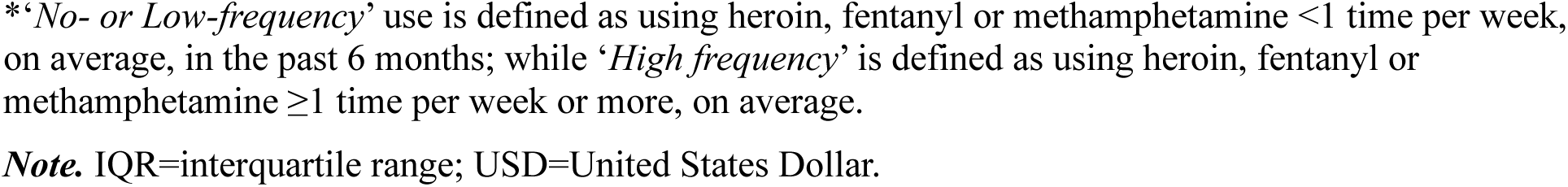
Substance use indicators, multilevel exposures of interest, and sociodemographic characteristics of 612 *La Frontera* participants at baseline, 2020-2021.

|  | **Total**<br>**(N=612; 100.00%)<sup>a</sup>** |
| --- | --- |
| **Substance use indicators\*** |  |
| **Injected heroin (past 6 months)** |  |
| No- to Low-frequency | 141 (23.0) |
| High frequency | 471 (77.0) |
| **Smoked heroin (past 6 months)** |  |
| No- to Low-frequency | 501 (81.9) |
| High frequency | 111 (18.1) |
| **Injected fentanyl (past 6 months)** |  |
| No- to Low-frequency | 516 (84.3) |
| High frequency | 96 (15.7) |
| **Smoked fentanyl (past 6 months)** |  |
| No- to Low-frequency | 543 (88.7) |
| High frequency | 69 (11.3) |
| **Injected methamphetamine (past 6 months)** |  |
| No- to Low-frequency | 393 (64.2) |
| High frequency | 219 (35.8) |
| **Smoked methamphetamine (past 6 months)** |  |
| No- to Low-frequency | 292 (47.7) |
| High frequency | 320 (52.3) |
| **Multilevel exposures** |  |
| **Location of residence** |  |
| City of Tijuana, Mexico | 202 (33.0) |
| San Diego County, United States | 410 (67.0) |
| **Housing status (past 6 months)** |  |
| Unsheltered homelessness | 265 (43.3) |
| Sheltered homelessness | 130 (21.2) |
| Not experiencing homelessness | 213 (34.8) |
| Missing | 4 (0.7) |
| **Received syringes from a syringe services program (past 6 months)** | 224 (36.6) |
| **Experienced an overdose (past 6 months)** | 97 (15.9) |
| Missing | 2 (0.3) |
| **Sociodemographic characteristics** |  |
| **Median age (IQR)** | 43.0 (35.0, 52.0) |
| **Assigned male sex at birth** | 453 (74.0) |
| **Hispanic/Latino/Mexican** | 440 (71.9) |
| **Monthly income < 500 USD** | 339 (55.4) |
| **Completed at least secondary school education** | 262 (42.8) |
| **Incarceration history of jail or prison** | 395 (64.5) |
| Missing | 1 (0.2) |

<sup>a</sup> Percentage (%) values may not add to 100% due to rounding.

### LTA model selection and baseline subgroup descriptions

#### Model selection

For each LTA model with two to five subgroups, the likelihood-ratio G^2^ statistic, degrees of freedom, AIC, and BIC are presented in Supplemental Table 3; p-values for the G^2^ statistic are not reported due to the large degrees of freedom and sparsity typical of high-dimensional contingency tables in LTA (42). Instead, we prioritized information criteria, where lower AIC and BIC values indicate improved fit relative to model complexity. Based on these criteria, we compared the four- and five-subgroup models in terms of parsimony, subgroup size, and interpretability. While the five-subgroup model had a lower AIC, it yielded smaller subgroup sizes and less interpretable patterns. In contrast, the four-subgroup model had the lowest BIC and produced more parsimonious and substantively meaningful subgroups with adequate sizes across time points. Based on this balance of fit and interpretability, we selected the four-subgroup model as our final model.

#### Baseline subgroup descriptions

We labeled the first subgroup *“Heroin-Methamphetamine Polyroute”* because it had high item-response probabilities for high-frequency heroin injecting (92%) and smoking (70%), and methamphetamine injecting (58%) and smoking (78%; Figure 1). We labeled the second subgroup *“Methamphetamine-dominant Smoking”* because it had a high item-response probability for high-frequency methamphetamine smoking (57%) and low item-response probabilities (<50%) for other high-frequency behaviors. We labeled the third subgroup *“Fentanyl-Methamphetamine Smoking”* because it had high item-response probabilities for high-frequency fentanyl (77%) and methamphetamine (69%) smoking, and low item-response probabilities for other high-frequency behaviors. We labeled the fourth subgroup *“Heroin-dominant Injecting”* because it had a high item-response probability for high-frequency heroin injecting (84%) and low item-response probabilities for other high-frequency behaviors.

**Figure 1.**
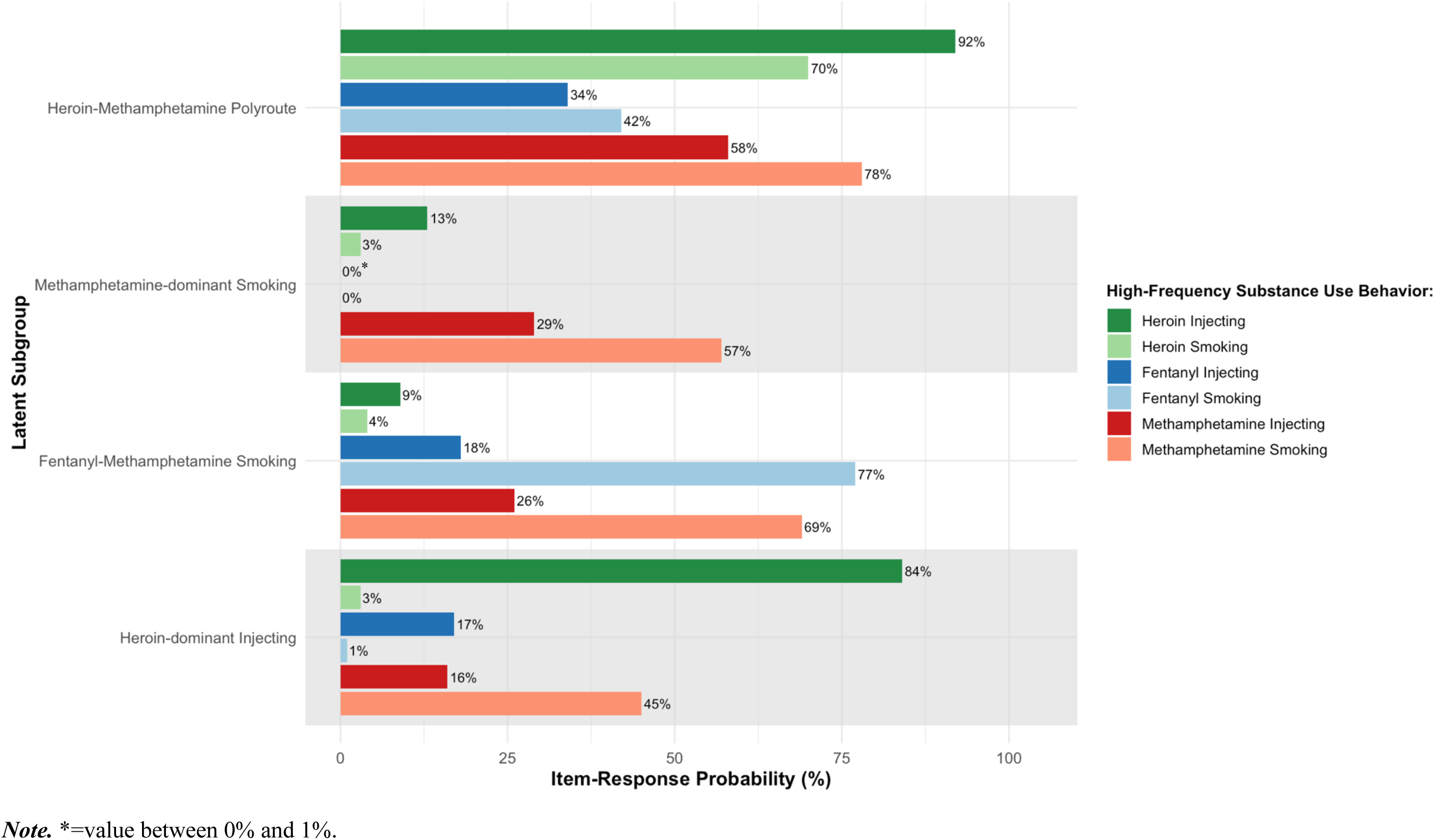
Item response probabilities for high-frequency substance use behaviors by baseline subgroups identified via latent transition analysis among 612 *La Frontera* participants, 2020–2021

#### Sensitivity analysis

At 18 months, 446 (73%) participants had not missed any follow-up visits, 55 (9%) missed one, 48 (8%) missed two, and 63 (10%) missed all three follow-up visits. After excluding participants missing at least one follow-up visit (n=166), the four-subgroup model continued to fit the data best, although the second subgroup had low item-response probabilities (<50%) for all high-frequency substance use behaviors. Given similar findings with respect to the number of subgroups and their descriptions, we proceeded with the full sample.

## Transition Analysis

### Overall Sample

#### Estimated prevalence of each subgroup over time

The *Heroin-dominant Injecting* subgroup was the most prevalent subgroup and its prevalence remained relatively stable over time (61% at baseline and 65% at 18 months; Figure 2). The prevalence of the *Methamphetamine-dominant Smoking* subgroup also remained stable over time (14% at baseline and 18 months), while the prevalence of the *Heroin-Methamphetamine Polyroute* subgroup decreased (22% at baseline to 0% at 18 months) and the prevalence of the *Fentanyl-Methamphetamine Smoking* subgroup increased (4% at baseline to 21% at 18 months).

**Figure 2.**
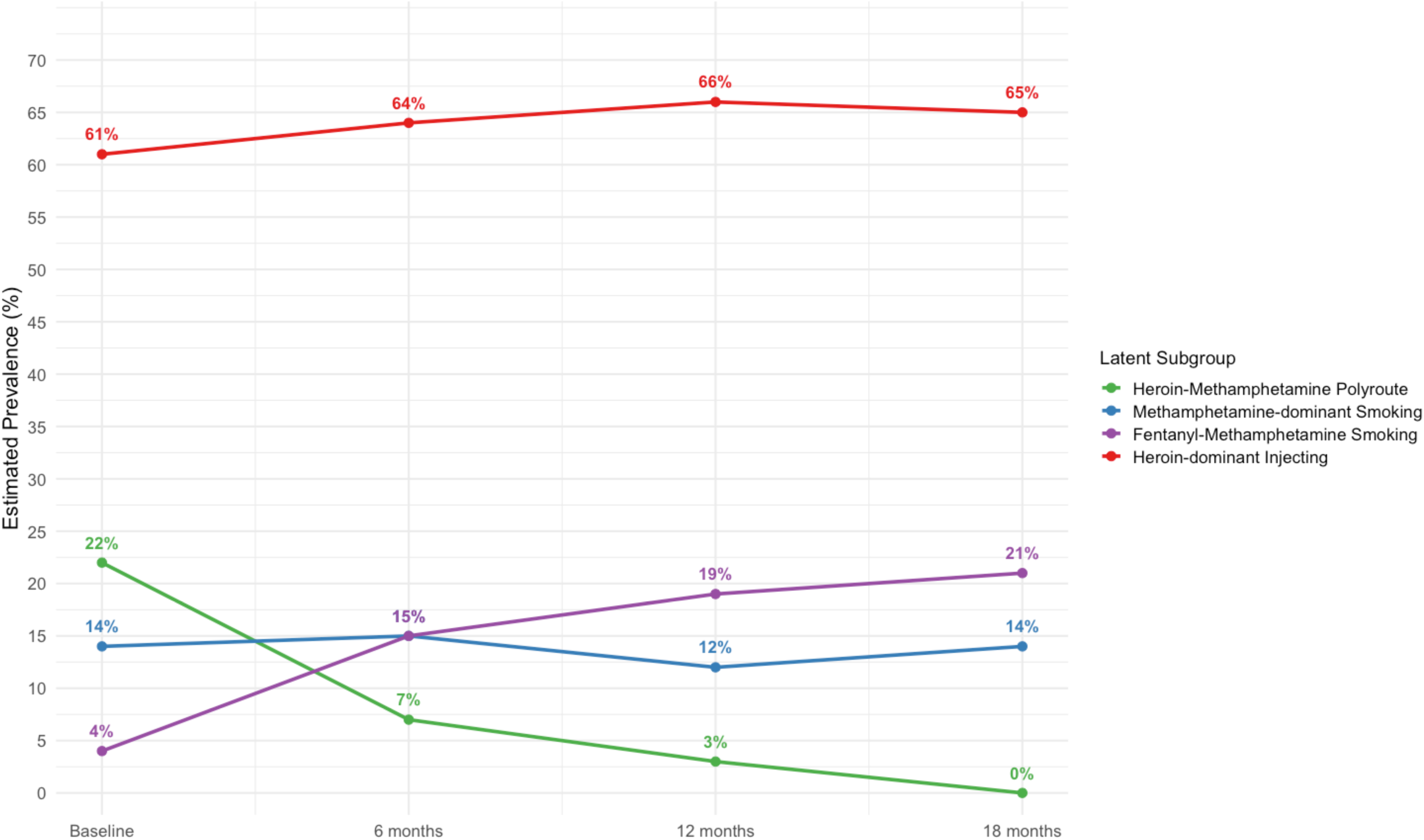
Estimated prevalence over time of latent subgroups identified via latent transition analysis among 612 *La Frontera* participants, 2020–2023

#### Transitions between subgroups over time

Transitions between subgroups over the 18-month follow-up period were common (Table 2). The *Heroin-Methamphetamine Polyroute* subgroup was least stable over time, with most participants transitioning from this subgroup elsewhere over the 18-month period. Across all six-month intervals of the follow-up period (baseline to six months, six to 12 months, and 12 to 18 months), a substantial proportion of participants in this subgroup transitioned to the *Heroin-dominant Injecting* (45%, 32%, and 45%, respectively) and *Fentanyl-Methamphetamine Smoking* subgroups (35%, 30% and 55%, respectively).

**Table 2.** Transition probabilities over time between latent subgroups identified via latent transition analysis among 612 *La Frontera* participants, 2020–2023.

| Transition time point | Initial subgroup | Follow-up subgroup |  |  |  |
| --- | --- | --- | --- | --- | --- |
|  |  | Heroin-Methamphetamine Polyroute | Methamphetamine-dominant Smoking | Fentanyl-Methamphetamine Smoking | Heroin-dominant Injecting |
| Baseline to 6 months | Heroin-Methamphetamine Polyroute | 11.2% | 8.7% | 35.3% | **44.9%** |
|  | Methamphetamine-dominant Smoking | 1.0% | **78.8%** | 0.0% | 20.2% |
|  | Fentanyl-Methamphetamine Smoking | 0.0% | 0.0% | **100.0%** | 0.0% |
|  | Heroin-dominant Injecting | 6.9% | 2.8% | 6.6% | **83.7%** |
| 6 months to 12 months | Heroin-Methamphetamine Polyroute | **37.9%** | 0.0% | 30.2% | 31.9% |
|  | Methamphetamine-dominant Smoking | 2.6% | **73.6%** | 0.6% | 23.2% |
|  | Fentanyl-Methamphetamine Smoking | 1.8% | 0.9% | **97.4%** | 0.0% |
|  | Heroin-dominant Injecting | 0.0% | 2.1% | 3.6% | **94.3%** |
| 12 months to 18 months | Heroin-Methamphetamine Polyroute | 0.0% | 0.0% | **55.0%** | 45.0% |
|  | Methamphetamine-dominant Smoking | 0.0% | **99.9%** | 0.1% | 0.0% |
|  | Fentanyl-Methamphetamine Smoking | 0.0% | 0.0% | **100.0%** | 0.0% |
|  | Heroin-dominant Injecting | 0.0% | 2.7% | 0.6% | **96.7%** |

*Note.* The largest transition probability per subgroup at each transition time point is bolded for ease of interpretation.

Participants in the *Methamphetamine-dominant Smoking* subgroup exhibited comparatively greater stability, where a substantial proportion of participants in this subgroup remained in the subgroup over time. However, during the first (baseline to six months) and second (six to 12 months) intervals of the follow-up period, transitions from this subgroup to the *Heroin-dominant Injecting* subgroup (20% and 23%, respectively) were also common.

The *Fentanyl-Methamphetamine Smoking* and *Heroin-dominant Injecting* subgroups were most stable over time, with nearly all participants in the *Fentanyl-Methamphetamine Smoking* (100%, 97%, and 100%, respectively) and *Heroin-dominant Injecting* (84%, 94%, and 97%, respectively) subgroups remaining in their respective subgroups over the follow-up period.

### Multiple-Groups LTA Results

Multiple-groups LTA model results revealed patterns similar to the sample overall; however, the magnitude, timing, and direction of transitions between subgroups differed meaningfully across all examined exposures, as noted below.

#### Housing status

The *Heroin-dominant Injecting* subgroup was the most prevalent subgroup over time in both strata and remained relatively stable (Supplemental Figure 1). However, its prevalence was consistently lower among participants who had experienced homelessness in the past six months at baseline (55% at baseline; 56% at 18 months) compared to those who had not (70% at baseline; 75% at 18 months). The *Fentanyl-Methamphetamine Smoking* subgroup increased in prevalence in both strata but increased to a greater extent among participants who had experienced homelessness (5% at baseline; 26% at 18 months) than among those who had not (1% at baseline; 11% at 18 months). The *Heroin-Methamphetamine Polyroute* subgroup declined to 0% in both strata over time, while the *Methamphetamine-dominant Smoking* subgroup remained relatively stable.

Transition probabilities further differentiated these strata (Supplemental Table 4). Among participants who had experienced homelessness, those in the *Heroin-Methamphetamine Polyroute* subgroup were most likely to transition to the *Fentanyl-Methamphetamine Smoking* subgroup at each transition time point (40%, 44%, and 56%, respectively). In contrast, among participants who had not experienced homelessness, transitions from the *Heroin-Methamphetamine Polyroute* subgroup varied over time: from baseline to six months, most transitioned to *Heroin-dominant Injecting* (66%); from six to 12 months, most remained in *Heroin-Methamphetamine Poly*route (46%); and from 12 to 18 months, most transitioned to *Fentanyl-Methamphetamine Smoking* (56%).

#### Receipt of syringes from an SSP

The *Heroin-dominant Injecting* subgroup was relatively stable and the most prevalent subgroup over time in both strata (Supplemental Figure 2); however, its prevalence was lower among participants who received syringes from an SSP in the past six months at baseline (50% at baseline and 18 months) than among those who did not (67% at baseline; 73% at 18 months). The *Fentanyl-Methamphetamine Smoking* subgroup increased in prevalence in both strata but was consistently higher and increased to a greater extent among participants who received syringes from an SSP (5% at baseline; 36% at 18 months) compared to those who did not (2% at baseline; 11% at 18 months). The *Heroin-Methamphetamine Polyroute* subgroup declined to 0% over time for both strata, while the prevalence of *Methamphetamine-dominant Smoking* remained relatively stable.

Transition probabilities were largely similar across strata (Supplemental Table 5), although two patterns likely contributed to differences in subgroup prevalences over time. First, participants who received syringes from an SSP had a lower probability of remaining in the *Heroin-dominant Injecting* subgroup during the first transition time point (64% from baseline to six months) compared to those who did not (89% from baseline to six months). Second, transitions from the *Heroin-Methamphetamine Polyroute* subgroup to *Fentanyl-Methamphetamine Smoking* were more common among participants who received syringes from an SSP during the first two transition time points (37% vs. 25% from baseline to six months; 34% vs. 23% from six to 12 months).

#### Overdose history

For participants who experienced an overdose in the past six months at baseline, there was substantial movement across subgroups over time (Supplemental Figure 3). From baseline to 18 months, there was a decline in the prevalence of the *Heroin-Methamphetamine Polyroute* subgroup (48% at baseline; 0% at 18 months), while the prevalence increased over time for the other three subgroups: 41% to 52% for *Heroin-dominant Injecting*; 6% to 36% for *Fentanyl-Methamphetamine Smoking*; and 5% to 12% for *Methamphetamine-dominant Smoking*. Meanwhile, for participants who had not experienced an overdose, the *Heroin-Methamphetamine Polyroute* subgroup also declined in prevalence over time (15% at baseline; 0% at 18 months), but only the *Fentanyl-Methamphetamine Smoking* subgroup (2% at baseline; 17% at 18 months) increased in prevalence over time.

From baseline to six months, participants who experienced an overdose in the *Heroin-Methamphetamine Polyroute* subgroup––the largest baseline subgroup for this stratum––had high probabilities of transitioning to the *Heroin-dominant Injecting* (42%) and the *Fentanyl-Methamphetamine Smoking* (36%) subgroups (Supplemental Table 6). However, from six to 12 months, participants who experienced an overdose in the *Heroin-Methamphetamine Polyroute* subgroup had a high probability of transitioning to the *Methamphetamine-dominant Smoking* (59%) subgroup as well as a high probability of remaining in the *Heroin-Methamphetamine Polyroute* subgroup (41%). Then, from 12 to 18 months, 100% of participants who experienced an overdose transitioned from the *Heroin-Methamphetamine Polyroute* subgroup to *Heroin-dominant Injecting*. Meanwhile, during the first two transition time points, participants who had not experienced an overdose in the *Heroin-Methamphetamine Polyroute* subgroup were most likely to transition to the *Heroin-dominant Injecting* (46% and 36%) and the *Fentanyl-Methamphetamine Smoking* (29% and 30%) subgroups, but they had a higher probability of transitioning to *Fentanyl-Methamphetamine Smoking* from 12 to 18 months (61%) than of transitioning to *Heroin-dominant Injecting* (35%) during that transition time point.

#### Location of Residence

The *Heroin-dominant Injecting* subgroup was less prevalent and relatively stable among San Diego participants (58% at baseline; 60% at 18 months) but more prevalent and increased over time among Tijuana participants (80% at baseline; 92% at 18 months; Supplemental Figure 4). In contrast, the *Fentanyl-Methamphetamine Smoking* subgroup was more prevalent in San Diego and increased over time (5% at baseline; 27% at 18 months), whereas it was nearly absent in Tijuana (<1% over time), which may explain why the multiple-groups LTA for ‘location of residence’ did not converge. The *Heroin-Methamphetamine Polyroute* subgroup declined in both locations (San Diego: 21% at baseline to 0% at 18 months; Tijuana: 11% at baseline to 0% at 18 months), while the *Methamphetamine-dominant Smoking* subgroup remained relatively stable.

## DISCUSSION

In this longitudinal cohort of PWID in the SDTBR, we identified four distinct subgroups of PWID with respect to their heroin, fentanyl, and methamphetamine injecting and smoking and transitions between those groups over time. These subgroups included a heroin and methamphetamine co-injecting and smoking (“polyroute”) subgroup, a methamphetamine-dominant smoking subgroup, a fentanyl and methamphetamine co-smoking subgroup, and a heroin-dominant injecting subgroup. Overall, membership in the high-frequency polyroute subgroup declined sharply, with many participants transitioning toward smoking-dominant patterns and less frequent polysubstance injection. However, increases in smoking-dominant use appeared to be driven primarily by growth in fentanyl-methamphetamine co-smoking behaviors, whereas methamphetamine-dominant smoking remained comparatively stable over time, suggesting that recent increases in smoking among PWID may have been more closely tied to opioid-involved smoking than to methamphetamine smoking alone.

The latent subgroups identified in this study represent meaningfully distinct substance use profiles for PWID using heroin, fentanyl, and methamphetamine between 2020 and 2023. Notably, the subgroup structure observed here differs from that identified in a recent LTA of PWID conducted on the U.S. East Coast (34), which largely distinguished subgroups by substance type and frequency rather than route of administration. A unique advantage of LTA is that it accounts for changes over time as well as baseline cohort composition. Therefore, it appears that recent declines in injecting––and rises in smoking––may have impacted our subgroup composition more drastically than the cohort on the East Coast, which corroborates nationally-representative data suggesting that PWID on the U.S. West Coast have decreased their injection drug use more substantially than those in other regions (5). This divergence from East Coast findings likely reflects region-specific contextual factors, including differences in drug market composition and harm reduction infrastructure, that likely influenced the magnitude of transitions from injecting to smoking across North America.

This is the first quantitative study, to our knowledge, to quantitatively assess the potential influence of experiencing an overdose on recent declines in injection drug use among PWID in North America. As a marker of distinct health-related harm from high-frequency polysubstance injection with opioids and stimulants in the fentanyl era (43, 44), participants who experienced an overdose were most likely to be in our ‘Heroin-Methamphetamine Polyroute’ subgroup at baseline. Then, after reporting a recent overdose, nearly all participants in this subgroup subsequently transitioned toward smoking-dominant patterns and less frequent polysubstance injection. These findings suggest the adoption of behavior perceived as harm reducing by PWID amidst rapid increases in morbidity and mortality caused by polysubstance use with opioids over the last decade (2, 43, 45). These findings corroborate earlier qualitative studies on transitions from injecting heroin to smoking fentanyl among PWID in San Francisco, CA (3), and British Columbia, Canada (8), that suggested transitions away from opioid injection reflect heightened awareness of overdose and other health harms from injecting fentanyl. Taken together, these findings indicate that PWID may intentionally respond to the downstream impacts of fentanyl adulteration by adopting behaviors perceived to decrease risk of immediate health threats––such as smoking. While a non-trivial amount of evidence suggests that transitions from injecting to smoking decreases risks for injection-related infections, it is less clear if this transition reduces risk of overdose (9, 44, 46). Nonetheless, future longitudinal research should examine whether population-level transitions from injecting to smoking translate into declines in overdose risk given the mounting evidence that PWID are transitioning away from injecting for this reason.

Importantly, we found that resources at the community level––including receiving syringes from an SSP––may have meaningfully shaped transitions toward fentanyl and methamphetamine co-smoking. SSPs serve as health hubs for PWID by providing low-barrier safer drug use equipment and non-stigmatizing services and referrals to primary care and addiction treatment (47). Existing work has shown that access to safer smoking equipment is associated with reduced injection frequency and increased smoking among PWID (48, 49). Even when accessed primarily for injection-related services like syringes, SSPs may facilitate transitions from injecting to smoking by legitimizing smoking as a harm-reducing practice. It is therefore plausible that, in the context of mounting injection-related harms, PWID accessing SSPs had the necessary knowledge and tools to facilitate transitions from injecting to smoking. However, additional qualitative evidence is necessary to further understand these dynamics.

While transitions from high-frequency heroin and methamphetamine polyroute use toward lower frequency polysubstance use and smoking appeared to vary by individual- and community-level exposures, the presence and distribution of these exposures were likely shaped by broader structural conditions. For example, larger increases in smoking-dominant behaviors among participants experiencing homelessness are consistent with evidence that PWID experience heightened surveillance and stigma relative to people who smoke drugs (3). Because injecting is more visible and time-intensive than smoking, transitions away from injecting may function as an adaptive strategy to reduce exposure to law enforcement and other forms of institutional control (8). Unsheltered homelessness, in particular, may intensify exposure to policing, displacement, and barriers to sterile syringe access, while simultaneously increasing reliance on lower-barrier, portable, and less conspicuous routes of drug administration like smoking (50, 51). Although we did not directly measure enforcement intensity or displacement in this study, intensified policing and involuntary displacement of people experiencing homelessness coincided with broader population-level shifts from injecting to smoking over the study period (52, 53), which may have impacted observed relationships in this sample.

Findings from our exploratory analyses by location of residence further underscore the role that place-based and structural vulnerabilities may play in shaping routes of administration. Importantly, fentanyl and methamphetamine co-smoking was nearly nonexistent in Tijuana, whereas participants in San Diego increased their prevalence of these behaviors five-fold. Meanwhile, membership in the heroin-dominant injecting subgroup remained large and increased modestly over time among participants residing in Tijuana. Although our data cannot directly identify the mechanisms underlying these differences, they may relate to variations in local drug supply composition (26), the timing and visibility of fentanyl penetration into the heroin market (18), and variation in access to harm reduction resources that may support route transitions, such as drug checking and safer smoking equipment (54). Together with our findings related to housing status and syringe access, these patterns suggest that structural vulnerability may have been a meaningful driver of transitions from injecting to smoking in the SDTBR. More broadly, these findings may help explain why declines in injecting may differ from eastern and western regions of North America and suggest that transitions from injecting to smoking are shaped by place-specific drug markets and service environments, not only individual preferences.

Several limitations should be considered when interpreting our findings. First, LTAs are sensitive to sample composition, and generalizability may be limited to settings with similar drug market dynamics, particularly other regions on the West Coast of North America. Similarly, cocaine use was excluded from analyses due to low prevalence in this cohort, which may limit comparability with studies conducted in regions where cocaine use is more common. Second, our LTA models assumed substance use indicator data were missing at random among participants who did not complete all follow-up visits. However, several characteristics differed between participants who did and did not complete all visits, suggesting this assumption may not fully hold and that selection bias is possible. Third, many of our measures relied on self-report and may be subject to recall or social desirability bias, although interviews were conducted by experienced bilingual staff trained to minimize such biases. Participants also may not have been aware of which drugs they used, which may bias results towards reporting heroin use instead of fentanyl use. Although, this discrepancy may have declined over time as fentanyl availability and awareness became more common in the SDTBR. Last, due to sample size constraints, we were unable to stratify our LTA by location of residence; which also contributed to the non-convergence of our multiple-groups model for this exposure. To address this limitation, we deterministically assigned participants to a latent subgroup according to their posterior probabilities. While this approach can introduce classification error, it is commonly used for exploratory examination of subgroup differences that would otherwise remain uncharacterized (38, 41). Despite these limitations, our study provides an important foundation for future work given the relative scarcity of longitudinal research examining transitions from injecting to smoking among PWID amidst unprecedented population-level changes over the last five years.

## CONCLUSION

This study provides the first longitudinal, multilevel quantitative evidence that recent population-level declines in injecting and increases in smoking among PWID are not uniform, but instead reflect heterogeneous trajectories that may be shaped by individual health experiences, access to harm reduction services, and structural vulnerability. These findings extend prior qualitative work by showing that harm-reduction logic surrounding smoking as a strategy to mitigate injection-related risks are reflected in behavioral transitions over time. Together, our results underscore that changes in routes of administration are best understood not as isolated individual decisions, but as adaptive responses to multilevel constraints and opportunities shaped by drug policy, service availability, and social marginalization. Future research should continue to examine whether these transitions translate to reductions in injection-related harms and overdose, and how harm reduction and policy responses can better support safer routes of drug use amid rapidly evolving drug supply changes.

## LIST OF ABBREVIATIONS

AIC: Akaike Information Criterion
BIC: Bayesian Information Criterion
EM: expectation-maximization
HCV: hepatitis C virus
HIV: human immunodeficiency virus
IQR: interquartile range
LTA: latent transition analysis
PWID: people who inject drugs
SDTBR: San Diego–Tijuana border region
SEM: social ecological model
SSP: syringe services program
U.S.: United States
USD: United States dollars

## DECLARATIONS OF INTEREST

### Ethics approval and consent to participate

The University of California, San Diego, and Universidad Xochicalco institutional review boards reviewed and approved all study activities. All participants provided written informed consent.

### Consent for publication

All authors have read the final draft of this manuscript and agreed to have it published.

### Availability of data and materials

Unidentified data is available upon reasonable request to Steffanie A. Strathdee.

### Authors’ contributions

- Conceptualization: WHE, ARB, SAS.
- Data curation: WHE, DA.
- Formal analysis: WHE, HAP.
- Investigation: AHV, CFV.
- Methodology: WHE, ELC, DA, HAP.
- Project administration: AHV, CFV.
- Resources: ARB, MGR, TLP, SAS.
- Supervision: ARB, SAS, HAP.
- Validation: WHE, DA, HAP.
- Writing – original draft: WHE, ARB, HAP.
- Writing – review and editing: ELC, DA, MGR, JRF, EVP, TLP, SAS.

## Data Availability

Unidentified data is available upon reasonable request to Steffanie A. Strathdee.

## Acknowledgements

The authors gratefully acknowledge the *La Frontera* participants who made this work possible, and the staff who play an integral role in data collection.

