## Supplemental Files for "Polysubstance Injection and Smoking Trajectories of Unregulated Drug Use in the San Diego-Tijuana Border Region: A Latent Transition Analysis"

**Supplemental Table 1. Self-reported substance use behaviors of 612 *La Frontera* participants at baseline, 2020-2021**

| **Variable** | **Total (n=612; 100.0%)^a^** |
| --- | --- |
| **Heroin use (past 6 months)** |  |
| Injected |  |
| Never | 77 (12.6) |
| One time per month or less | 24 (3.9) |
| 2–3 days per month | 21 (3.4) |
| One time per week | 19 (3.1) |
| 2–3 days per week | 43 (7.0) |
| 4–6 days per week | 9 (1.5) |
| One time per day | 28 (4.6) |
| 2–3 times per day | 193 (31.5) |
| 4 times or more per day | 198 (32.4) |
| Smoked |  |
| Never | 444 (72.6) |
| One time per month or less | 34 (5.6) |
| 2–3 days per month | 13 (2.1) |
| One time per week | 10 (1.6) |
| 2–3 days per week | 28 (4.6) |
| 4–6 days per week | 5 (0.8) |
| One time per day | 14 (2.3) |
| 2–3 times per day | 39 (6.4) |
| 4 times or more per day | 25 (4.1) |
| **Fentanyl use (past 6 months)** |  |
| Injected |  |
| Never | 453 (74.0) |
| One time per month or less | 42 (6.9) |
| 2–3 days per month | 13 (2.1) |
| One time per week | 8 (1.3) |
| 2–3 days per week | 20 (3.3) |
| 4–6 days per week | 6 (1.0) |
| One time per day | 11 (1.8) |
| 2–3 times per day | 23 (3.8) |
| 4 times or more per day | 36 (5.9) |
| Smoked |  |
| Never | 494 (80.7) |
| One time per month or less | 30 (4.9) |
| 2–3 days per month | 11 (1.8) |
| One time per week | 8 (1.3) |
| 2–3 days per week | 8 (1.3) |
| 4–6 days per week | 3 (0.5) |
| One time per day | 11 (1.8) |
| 2–3 times per day | 17 (2.8) |
| 4 times or more per day | 30 (4.9) |
| **Methamphetamine use (past 6 months)** |  |
| Injected |  |
| Never | 291 (47.6) |
| One time per month or less | 56 (9.2) |
| 2–3 days per month | 23 (3.8) |
| One time per week | 23 (3.8) |
| 2–3 days per week | 30 (4.9) |
| 4–6 days per week | 2 (0.3) |
| One time per day | 24 (3.9) |
| 2–3 times per day | 67 (11.0) |
| 4 times or more per day | 96 (15.7) |
| Smoked |  |
| Never | 218 (35.6) |
| One time per month or less | 38 (6.2) |
| 2–3 days per month | 19 (3.1) |
| One time per week | 17 (2.8) |
| 2–3 days per week | 55 (9.0) |
| 4–6 days per week | 9 (1.5) |
| One time per day | 47 (7.7) |
| 2–3 times per day | 114 (18.6) |
| 4 times or more per day | 95 (15.5) |

^a^ Percentage (%) values may not add to 100% due to rounding.

**Supplemental Table 2. Substance use indicators, multilevel exposures of interest, and sociodemographic characteristics of 612 *La Frontera* participants at baseline by loss to follow-up status, 2020–2023**

|  | **Missed at least one study visit**  **(n=166; 27.1%)^a^** | **Did not miss any study visits**  **(n=446; 72.9%)^a^** |
| --- | --- | --- |
| **Substance use indicators*** |  |  |
| **Injected heroin (past 6 months)** |  |  |
| No- to Low-frequency | 54 (32.5) | 87 (19.5) |
| High frequency | 112 (67.5) | 359 (80.5) |
| **Smoked heroin (past 6 months)** |  |  |
| No- to Low-frequency | 121 (72.9) | 380 (85.2) |
| High frequency | 45 (27.1) | 66 (14.8) |
| **Injected fentanyl (past 6 months)** |  |  |
| No- to Low-frequency | 141 (84.9) | 375 (84.1) |
| High frequency | 25 (15.1) | 71 (15.9) |
| **Smoked fentanyl (past 6 months)** |  |  |
| No- to Low-frequency | 129 (77.7) | 414 (92.8) |
| High frequency | 37 (22.3) | 32 (7.2) |
| **Injected methamphetamine (past 6 months)** |  |  |
| No- to Low-frequency | 107 (64.5) | 286 (64.1) |
| High frequency | 59 (35.5) | 160 (35.9) |
| **Smoked methamphetamine (past 6 months)** |  |  |
| No- to Low-frequency | 57 (34.3) | 235 (52.7) |
| High frequency | 109 (65.7) | 211 (47.3) |
| **Multilevel exposures** |  |  |
| **Location of residence** |  |  |
| City of Tijuana, Mexico | 12 (7.2) | 190 (42.6) |
| San Diego County, United States | 154 (92.8) | 256 (57.4) |
| **Housing status (past 6 months)** |  |  |
| Unsheltered homelessness | 96 (57.8) | 169 (37.9) |
| Sheltered homelessness | 30 (18.1) | 100 (22.4) |
| Not experiencing homelessness | 39 (23.5) | 174 (39.0) |
| Missing | 3 (0.7) | 1 (0.6) |
| **Received syringes from a syringe services program (past 6 months)** | 78 (47.0) | 146 (32.7) |
| **Experienced an overdose (past 6 months)** | 61 (13.7) | 36 (21.7) |
| Missing | 2 (1.2) | 0 (0.0) |
| **Sociodemographic characteristics** |  |  |
| **Median age (IQR)** |  |  |
| **Assigned male sex at birth** | 125 (75.3) | 328 (73.5) |
| **Hispanic/Latino/Mexican** | 77 (46.4) | 363 (81.4) |
| **Monthly income <500 USD** | 66 (39.8) | 273 (61.2) |
| **Completed at least secondary school education** | 33 (19.9) | 229 (51.4) |
| **Incarceration history of jail or prison** | 116 (69.9) | 279 (62.6) |
| Missing | 1 (0.60) | 0 (0.0) |

^a^ Percentage (%) values may not add to 100% due to rounding.

*‘*No- or Low-frequency*’ use is defined as using heroin, fentanyl or methamphetamine <1 time per week, on average, in the past 6 months; while ‘*High frequency*’ is defined as using heroin, fentanyl or methamphetamine ≥1 time per week or more, on average.

***Note.*** IQR=interquartile range; USD=United States Dollar.

Supplemental Table 3. Fit statistics from latent transition analysis models fit to substance use indicator data from 612 *La Frontera* participants, 2020-2023

|  | **Fit Statistics** | | | |
| --- | --- | --- | --- | --- |
| **Number of Subgroups** | **Likelihood-Ratio G^2^** | **Degrees of Freedom** | **AIC** | **BIC** |
| 2 | 5002.16 | 16,777,196 | 5040.16 | 5124.08 |
| 3 | 4629.56 | 16,777,177 | 4705.25 | 4873.08 |
| **4** | **4458.79** | **16,777,152** | **4584.79** | **4863.04** |
| 5 | 4303.46 | 16,777,121 | 4491.46 | 4906.64 |

***Note.*** AIC; Akaike information criteria; BIC, Bayesian Information Criteria; bold font indicates the selected model.

**Supplemental Figure 1. Estimated prevalence of latent subgroups over time identified via latent transition analysis among 612 *La Frontera* participants by past six-month housing status, 2020–2023**

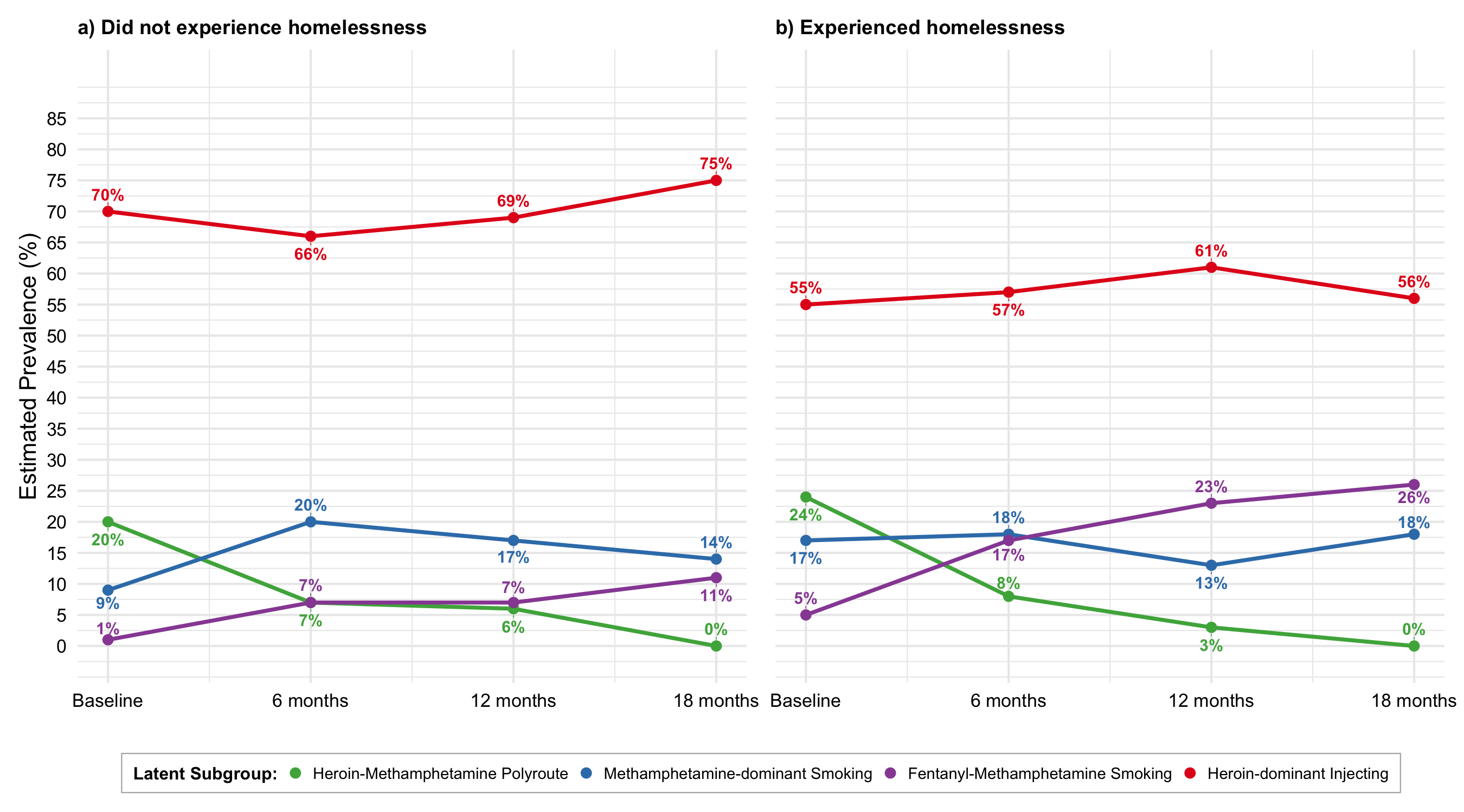

*****

***Note.*** *=value between 0% and 1%.

**Supplemental Table 4. Transition probabilities of latent subgroups over time identified via latent transition analysis among 612 *La Frontera* participants by past six-month housing status, 2020–2023**

| **Transition time point** | **Initial subgroup** | **Follow-up subgroup** | | | | | | | |
| --- | --- | --- | --- | --- | --- | --- | --- | --- | --- |
|  |  | **Did not experience homelessness** | | | | **Experienced homelessness** | | | |
|  |  | Heroin-Meth Polyroute | Meth-dominant Smoking | Fentanyl-Meth Smoking | Heroin-dominant Injecting | Heroin-Meth Polyroute | Meth-dominant Smoking | Fentanyl-Meth Smoking | Heroin-dominant Injecting |
| **Baseline to 6 months** | Heroin-Meth Polyroute | 8.7% | 8.5% | 17.2% | **65.6%** | 14.7% | 15.6% | **39.5%** | 30.2% |
|  | Meth-dominant Smoking | 10.7% | **83.2%** | 0.0% | 6.1% | 1.1% | **75.0%** | 1.5% | 22.4% |
|  | Fentanyl-Meth Smoking | 0.0% | 0.0% | **100.0%** | 0.0% | 0.0% | 0.0% | **100.0%** | 0.0% |
|  | Heroin-dominant Injecting | 6.1% | 14.6% | 4.7% | **74.6%** | 7.5% | 3.0% | 6.2% | **83.3%** |
| **6 months to 12 months** | Heroin-Meth Polyroute | **45.9%** | 18.6% | 6.2% | 29.4% | 27.8% | 0.0% | **44.1%** | 28.1% |
|  | Meth-dominant Smoking | 1.7% | **64.3%** | 0.0% | 34.0% | 1.9% | **69.2%** | 3.7% | 25.3% |
|  | Fentanyl-Meth Smoking | 0.0% | 5.6% | **91.6%** | 2.9% | 2.4% | 0.0% | **97.6%** | 0.0% |
|  | Heroin-dominant Injecting | 4.0% | 4.7% | 0.0% | **91.4%** | 0.0% | 0.6% | 3.7% | **95.7%** |
| **12 months to 18 months** | Heroin-Meth Polyroute | 0.0% | 0.0% | **55.9%** | 44.1% | 12.1% | 8.2% | **55.7%** | 24.0% |
|  | Meth-dominant Smoking | 0.0% | **82.4%** | 3.3% | 14.3% | 0.0% | **90.5%** | 9.5% | 0.0% |
|  | Fentanyl-Meth Smoking | 0.0% | 0.0% | **100.0%** | 0.0% | 0.0% | 3.4% | **96.6%** | 0.0% |
|  | Heroin-dominant Injecting | 0.0% | 0.0% | 0.0% | **100.0%** | 0.0% | 8.1% | 0.8% | **91.0%** |

***Note.*** The largest transition probability per subgroup at each transition time point is bolded for ease of interpretation.

### **Supplemental Figure 2. Estimated prevalence of latent subgroups over time identified via latent transition analysis among 612 *La Frontera* participants by past six-month receipt of syringes from a syringe services program, 2020–2023**

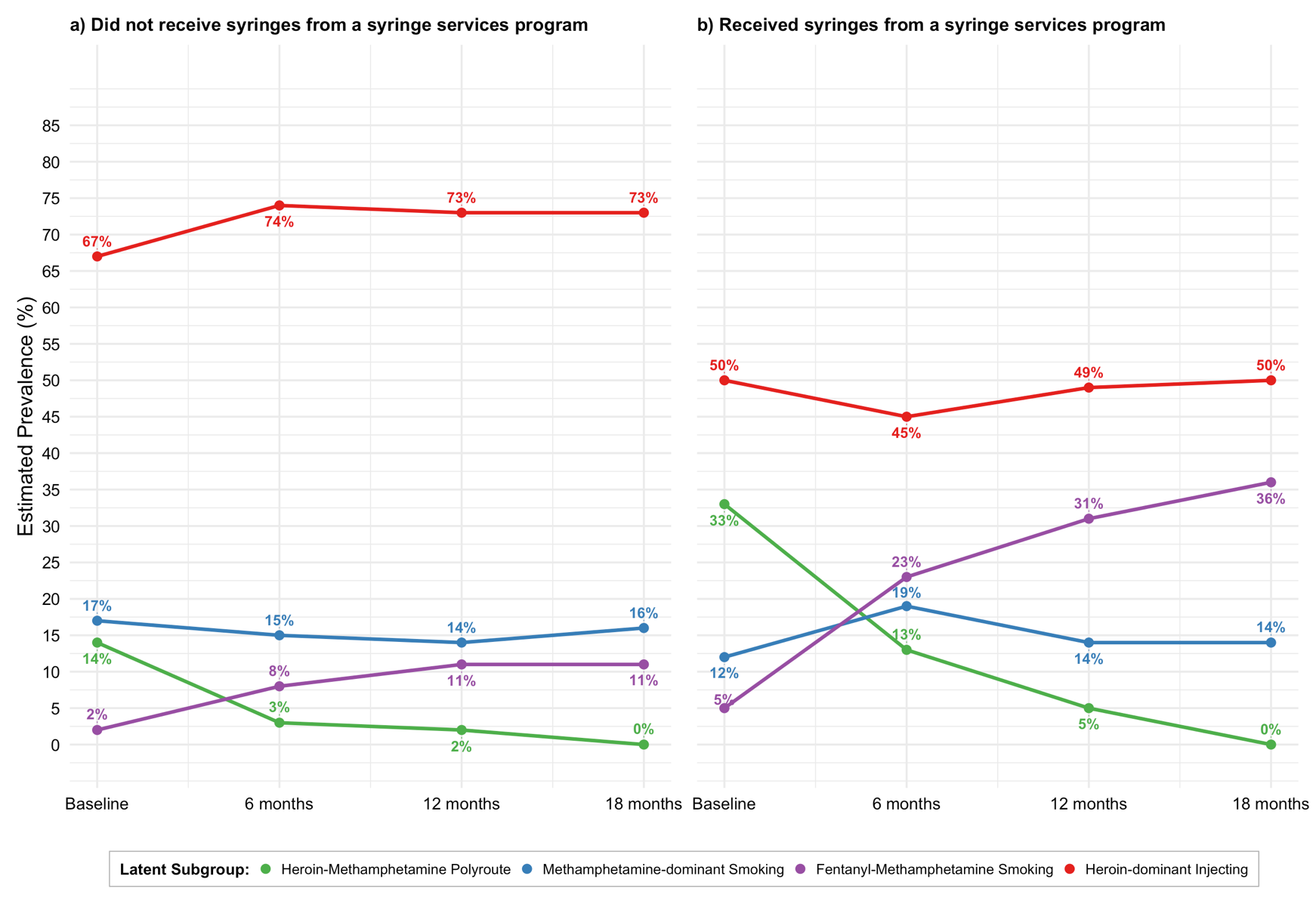

*****

***Note.*** *=value between 0% and 1%.

**Supplemental Table 5. Transition probabilities of latent subgroups over time identified via latent transition analysis among 612 *La Frontera* participants by past six-month receipt of syringes from a syringe services program (SSP), 2020–2023**

| **Transition time point** | **Initial subgroup** | **Follow-up subgroup** | | | | | | | |
| --- | --- | --- | --- | --- | --- | --- | --- | --- | --- |
|  |  | **Did not receive syringes from an SSP** | | | | **Received syringes from an SSP** | | | |
|  |  | Heroin-Meth Polyroute | Meth-dominant Smoking | Fentanyl-Meth Smoking | Heroin-dominant Injecting | Heroin-Meth Polyroute | Meth-dominant Smoking | Fentanyl-Meth Smoking | Heroin-dominant Injecting |
| **Baseline to 6 months** | Heroin-Meth Polyroute | 4.4% | 13.8% | 25.2% | **56.6%** | 14.2% | 11.0% | 36.7% | **38.2%** |
|  | Meth-dominant Smoking | 5.0% | **60.9%** | 0.0% | 34.1% | 0.0% | **98.6%** | 1.4% | 0.0% |
|  | Fentanyl-Meth Smoking | 0.0% | 0.0% | **100.0%** | 0.0% | 0.0% | 0.0% | **100.0%** | 0.0% |
|  | Heroin-dominant Injecting | 2.4% | 3.9% | 4.5% | **89.2%** | 17.2% | 8.1% | 10.4% | **64.2%** |
| **6 months to 12 months** | Heroin-Meth Polyroute | **34.5%** | 33.5% | 22.8% | 9.2% | **36.3%** | 0.0% | 34.2% | 29.5% |
|  | Meth-dominant Smoking | 4.7% | **68.4%** | 0.0% | 27.0% | 0.0% | **71.6%** | 4.8% | 23.6% |
|  | Fentanyl-Meth Smoking | 0.0% | 0.0% | **100.0%** | 0.0% | 2.5% | 1.9% | **95.7%** | 0.0% |
|  | Heroin-dominant Injecting | 0.0% | 4.5% | 2.1% | **93.5%** | 0.0% | 0.0% | 9.4% | **90.7%** |
| **12 months to 18 months** | Heroin-Meth Polyroute | 0.0% | 0.0% | **50.5%** | 49.5% | 8.6% | 7.9% | 41.2% | **42.3%** |
|  | Meth-dominant Smoking | 0.0% | **99.4%** | 0.0% | 0.7% | 0.0% | **65.5%** | 18.1% | 16.4% |
|  | Fentanyl-Meth Smoking | 0.0% | 0.0% | **91.7%** | 8.3% | 0.0% | 0.0% | **100.0%** | 0.0% |
|  | Heroin-dominant Injecting | 0.0% | 2.2% | 0.9% | **96.9%** | 0.0% | 8.1% | 0.0% | **91.9%** |

***Note.*** The largest transition probability per subgroup at each transition time point is bolded for ease of interpretation.

### **Supplemental Figure 3. Estimated prevalence of latent subgroups over time identified via latent transition analysis among 612 *La Frontera* participants by past six-month overdose history, 2020–2023**

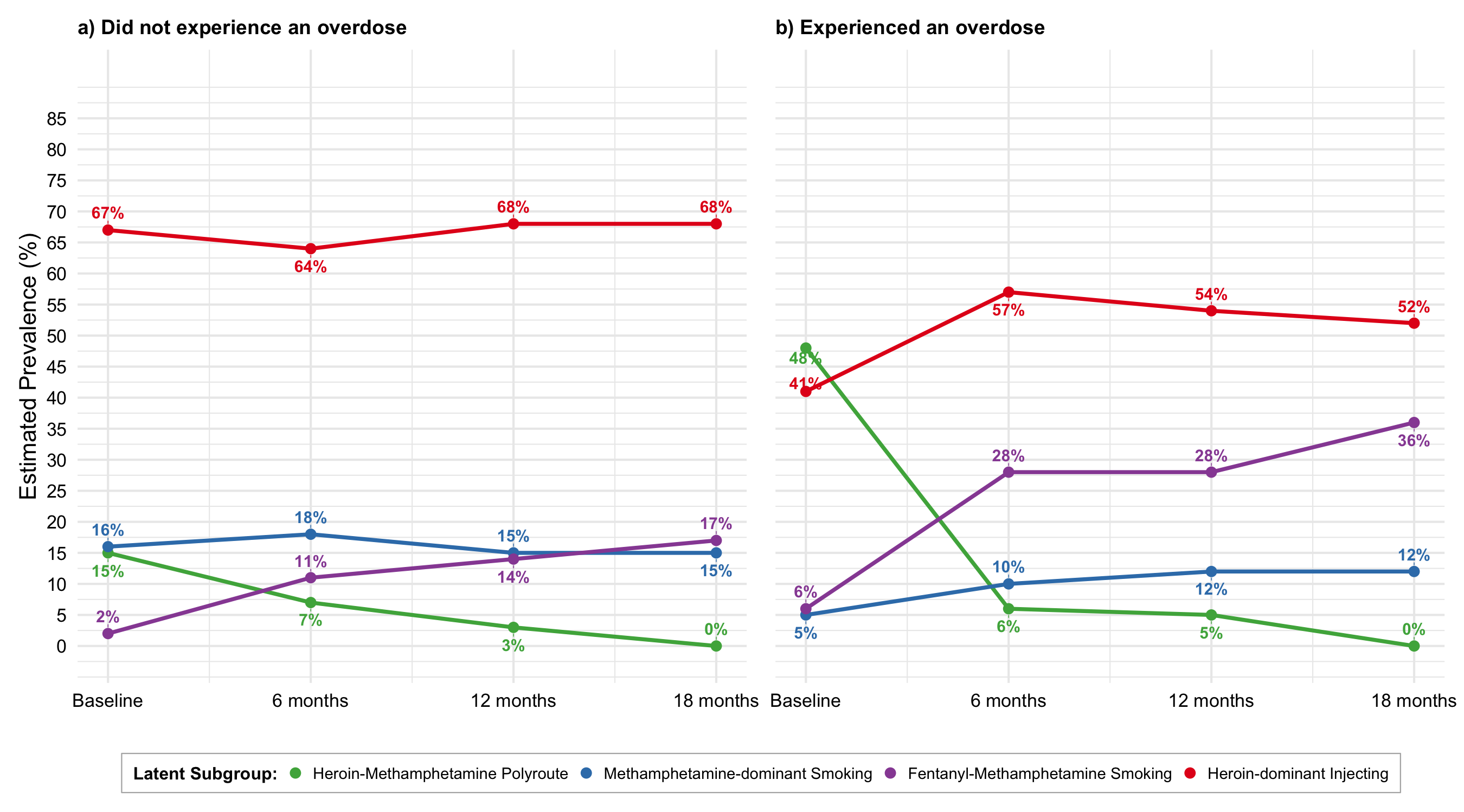

**Supplemental Table 6. Transition probabilities of latent subgroups over time identified via latent transition analysis among 612 *La Frontera* participants by past six-month overdose history, 2020-2023**

| **Transition time point** | **Initial subgroup** | **Follow-up subgroup** | | | | | | | |
| --- | --- | --- | --- | --- | --- | --- | --- | --- | --- |
|  |  | **Did not experience an overdose** | | | | **Experienced an overdose** | | | |
|  |  | Heroin-Meth Polyroute | Meth-dominant Smoking | Fentanyl-Meth Smoking | Heroin-dominant Injecting | Heroin-Meth Polyroute | Meth-dominant Smoking | Fentanyl-Meth Smoking | Heroin-dominant Injecting |
| **Baseline to 6 months** | Heroin-Meth Polyroute | 12.1% | 13.1% | 28.7% | **46.0%** | 12.0% | 10.2% | 35.7% | **42.0%** |
|  | Meth-dominant Smoking | 1.8% | **72.2%** | 0.0% | 25.9% | 0.0% | 25.5% | **74.5%** | 0.0% |
|  | Fentanyl-Meth Smoking | 0.0% | 0.0% | **100.0%** | 0.0% | 0.0% | 0.0% | **100.0%** | 0.0% |
|  | Heroin-dominant Injecting | 7.4% | 7.4% | 6.1% | **79.2%** | 0.0% | 8.3% | 1.7% | **90.1%** |
| **6 months to 12 months** | Heroin-Meth Polyroute | 34.8% | 0.0% | 29.7% | **35.5%** | 41.2% | **58.8%** | 0.0% | 0.0% |
|  | Meth-dominant Smoking | 2.7% | **72.3%** | 0.0% | 25.0% | 0.0% | 0.0% | **50.4%** | 49.6% |
|  | Fentanyl-Meth Smoking | 0.0% | 0.9% | **99.1%** | 0.0% | 5.6% | 32.8% | **61.6%** | 0.0% |
|  | Heroin-dominant Injecting | 0.0% | 3.0% | 2.4% | **94.6%** | 2.5% | 0.0% | 11.1% | **86.4%** |
| **12 months to 18 months** | Heroin-Meth Polyroute | 0.0% | 4.1% | **60.7%** | 35.3% | 0.0% | 0.0% | 0.0% | **100.0%** |
|  | Meth-dominant Smoking | 0.0% | **83.2%** | 8.4% | 8.4% | 0.0% | 36.2% | **63.8%** | 0.0% |
|  | Fentanyl-Meth Smoking | 0.0% | 2.7% | **96.0%** | 1.3% | 0.0% | 0.0% | **100.0%** | 0.0% |
|  | Heroin-dominant Injecting | 0.0% | 2.9% | 0.6% | **96.5%** | 0.0% | 14.2% | 0.0% | **85.8%** |

***Note.*** The largest transition probability per subgroup at each transition time point is bolded for ease of interpretation.

### **Supplemental Figure 4. Prevalence of latent subgroups over time among 612 *La Frontera* participants by location of residence, 2020–2023**

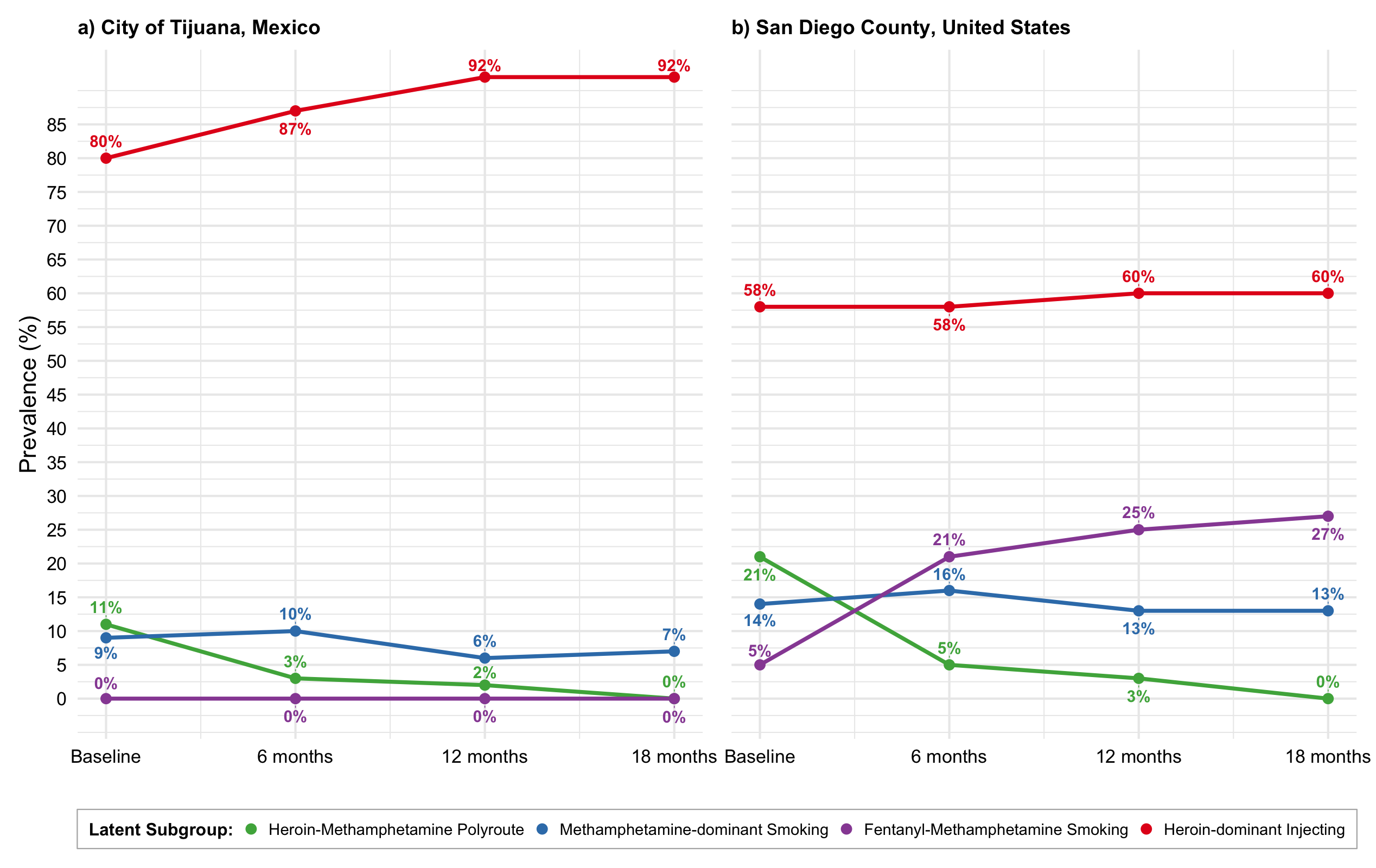

*****

*****

***Note.*** *=value between 0% and 1%.
